# Socioeconomic Position, Kidney Outcomes, and Mortality: A Longitudinal Cohort Study from the All of Us Research Program

**DOI:** 10.64898/2026.11.11.26367876

**Authors:** Atalay Demiray, Ibrahim Gulmaliyev, Mehmet Ali Enlice, Sidar Copur, Metehan Berkkan, Francesca Mallamaci, Carmine Zoccali, Mehmet Kanbay

## Abstract

**Background:** Socioeconomic position is associated with kidney disease, but it remains unclear whether these differences are greater for worsening kidney function than for a first recorded diagnosis. We examined how household income, education, and neighborhood deprivation relate to mortality and kidney outcomes.

**Methods and findings:** We conducted a retrospective cohort study involving 218,098 individuals using data from the All of Us Research Program, with follow-up to January 1, 2025. Primary outcomes involved all-cause mortality and sustained 40% estimated glomerular filtration rate decline. Secondary outcomes were incident recorded chronic kidney disease (CKD) diagnosis, incident recorded CKD category G3a or worse, and kidney failure. We also evaluated income inequalities and estimated standardized 3-year mortality risks for each baseline kidney state.

Median follow-up was 3.62 years for mortality and 2.76 years for kidney outcomes. Among 218,098 individuals, 4,777 deaths, 6,423 sustained declines, 7,870 incident recorded chronic kidney disease diagnoses, 4,716 incident category G3a-or-worse events, and 1,799 kidney failure events were observed. Adjusted hazard ratios (95% CIs) for household income below $25,000 compared with $200,000 or more were 2.68 (2.31–3.12), 3.57 (3.08–4.13), 2.13 (1.90–2.38), 2.15 (1.86–2.49), and 3.08 (2.30–4.11), respectively. Relative index of inequality estimates were approximately twice as large for sustained decline and kidney failure than for recorded CKD diagnosis. Within baseline kidney states, standardized 3-year absolute mortality differences by income did not differ significantly across states. The main limitations are the observational design, baseline creatinine measured up to a year after the survey, and outcome ascertainment from health records that may differ by income.

**Conclusions:** Lower income was associated with higher mortality and greater risk of all kidney outcomes. Education and neighborhood deprivation showed different patterns of association, which supports considering individual and neighborhood circumstances together.

## Introduction

Health inequities are systematic, avoidable, and unfair differences in health outcomes and access to healthcare between social groups, rooted in unequal social, economic, and environmental conditions (1). Globally, an estimated 2.1 billion people, which represents 26% of the global population, experienced financial hardship from out-of-pocket healthcare spending in 2022. Approximately 75% of people in the poorest groups faced financial hardship from healthcare costs compared with fewer than 4% in the richest groups (2). These inequalities are particularly relevant to kidney health (3, 4). Among 14,086 participants in the Atherosclerosis Risk in Communities (ARIC) study with a median follow-up duration of 23 years, lower income was associated with a 15% steeper decline in estimated glomerular filtration rate (eGFR) and a higher risk of kidney failure (adjusted HR, 1.50; 95% CI, 1.14–1.98) compared with high income (5). Similarly, among 5.6 million Japanese working adults, the lowest income decile, compared with the highest, was associated with rapid kidney function decline (adjusted OR, 1.70; 95% CI, 1.67-1.73) and higher risk of renal replacement therapy initiation (adjusted HR, 1.65; 95% CI, 1.47-1.86) (6). In the Scottish GLOMMS-CORE study, people with early kidney disease living in unemployed households had higher mortality than those in professional households (adjusted HR, 1.77; 95% CI, 1.60-1.96) (7). Both household and neighborhood deprivation were associated with higher mortality, with the poorest outcomes among people experiencing both. These findings highlight the role of socioeconomic inequalities within adverse renal outcomes and mortality. Nevertheless, limited evidence exists about the association of income inequalities in recorded chronic kidney disease and sustained kidney function decline. We therefore conducted a large-scale cohort study using the All of Us Research Program Controlled Tier Dataset examine the role of socioeconomic inequalities across adverse renal outcomes and mortality.

## Materials and Methods

### Study design and population

We conducted a retrospective cohort study using the All of Us Research Program Controlled Tier Dataset (version 9, C2025Q4R6) which enrolls adults across the United States with deliberate recruitment of underrepresented populations in biomedical research (8). Participants provided informed consent, and the All of Us program protocol was approved by its Institutional Review Board (protocol 11-01139). The Yale University Institutional Review Board determined that this analysis was not human subjects research (protocol 2000043320; July 14, 2026). Counts of 1 to 20 were suppressed in accordance with All of Us policy. This study followed the STROBE reporting guideline (9).

The study included participants aged 18 years or older with linked electronic health record data, determined baseline kidney state, positive mortality follow-up after the index date, and provided substantive responses to both household income and educational attainment. The index date was the earliest response to either socioeconomic item and could precede the other response. Participants without a creatinine measurement from which estimated glomerular filtration rate could be calculated were eligible only if kidney failure or kidney transplantation could be identified. Missing responses, including “Skip” and “Prefer not to answer,” were excluded.

### Socioeconomic measures

Household income and educational attainment were obtained from participant surveys. Annual household income was categorized as <$25,000, $25,000–49,999, $50,000–99,999, $100,000– 199,999, or ≥$200,000. Educational attainment was categorized as less than high school, high school or equivalent, some college, or college graduate or above. Income was not adjusted for household size. Neighborhood deprivation was the program-supplied index based on 3-digit ZIP code and was categorized into tertiles using the otherwise-eligible source cohort. Neighborhood deprivation values that were unavailable were coded separately as a different category. Income, education, and neighborhood deprivation were included in the primary models.

### Baseline kidney state

Estimated glomerular filtration rate was calculated from serum creatinine using the race-free 2021 CKD-EPI equation (10). Baseline creatinine was the measurement closest to the index within 365 days before or after that date. Creatinine values between 0.1 and 20 were retained and treated as mg/dL; recorded units were not used to filter or convert values. Participants without kidney failure or transplantation were classified into GFR categories G1 (≥90), G2 (60–89), G3a (45–59), G3b (30–44), G4 (15–29), or G5 (<15 mL/min/1.73 m²). Kidney failure and transplantation were classified as separate baseline states, with transplantation taking precedence. Kidney failure was identified from diagnostic codes indicating chronic kidney failure or dialysis dependence or from dialysis records on at least 2 dates separated by at least 90 days. Kidney-failure qualification used the available longitudinal record, with onset assigned to the earliest dialysis/ESKD record; later evidence could retrospectively qualify a date before index. Kidney transplantation was identified from status or procedure records. Detailed information is provided in Supplementary Methods.

### Outcomes and follow-up

The primary outcomes were all-cause mortality and a sustained 40% decline in eGFR. Decline required an eGFR ≤60% of baseline after the baseline measurement, confirmed by another qualifying value at least 30 days later. The event was dated to the first qualifying measurement. Qualifying low eGFR values did not need to be consecutive, and the confirmation period was not explicitly restricted by death or the administrative end of follow-up. Prevalent recorded CKD was identified when the assigned first-record date was on or before the index date, although confirmation of CKD could occur after the index date. Further details are provided in Supplementary Methods. Secondary outcomes were a first recorded diagnosis of chronic kidney disease (CKD), first recorded CKD category G3a or worse, and kidney failure. Recorded CKD outcomes required one inpatient diagnosis or qualifying diagnoses on at least two different dates, with the event dated to the first record. The kidney-failure outcome was the first qualifying dialysis or ESKD record, as defined above, or kidney transplantation, whichever occurred first. Participants with recorded CKD, kidney failure, or transplantation at baseline were excluded from the recorded-diagnosis analyses. Participants with baseline kidney failure or transplantation were excluded from the decline and kidney-failure analyses. The decline analysis also required a baseline eGFR estimate. Each analysis required positive follow-up. For kidney outcomes, follow-up ended at qualifying outcome, death, last qualifying clinical contact, or January 1, 2025, whichever occurred first. Mortality follow-up ended at death or January 1, 2025, without censoring at the last EHR contact. Deaths were identified from the program’s death records, using the earliest recorded date for each participant.

### Statistical analysis

Adjusted hazard ratios and 95% confidence intervals (CIs) were estimated using Cox models with the Efron method of handling ties. Household income, education, and neighborhood deprivation were included in primary Cox models as categorical variables. Models were adjusted for age, sex, race, and ethnicity. Race and ethnicity were self-reported in the program’s basics survey and were included as social categories to account for social patterning of socioeconomic position, kidney disease, and care; the categories are listed in the Supplementary Methods. Age was modeled linearly. Remaining variables were treated as categorical. Reference groups were income ≥$200,000, college graduate or above, and the least deprived neighborhood tertile. For nonfatal outcomes, cause-specific models censored participants at death. The proportional hazards assumption was assessed with scaled Schoenfeld residual tests. Follow-up was split at 2 years, and income associations were estimated separately within each period with the same covariate set. When the assumption was not met, overall HRs were interpreted as summaries of the associations over follow-up. Income inequalities were also summarized using the relative index of inequality (RII) and slope index of inequality (SII) (11). For each outcome cohort, income groups were ranked from highest to lowest. Each group received a ridit score equal to the proportion of participants in higher-income groups plus half the proportion in that group. RII compared the extremes of income rank using Cox models adjusted for sex, age, race, and ethnicity. The RII is the hazard ratio for a one-unit change in ridit score, the modeled contrast between the lowest- and highest-income extremes of the distribution; values above 1 indicate higher hazard with lower income. The indices summarize the total income gradient and were therefore not adjusted for education or neighborhood deprivation. SII used population-weighted regression of the five crude income-category rates on ridit scores and was expressed per 1000 person-years. Further details are provided in Supplementary Methods. In exploratory analyses, we estimated the income trend per $10,000 within subgroups defined by race, sex, age, diabetes, and baseline kidney state, and the categorical socioeconomic associations with mortality within sex, race, and neighborhood-deprivation strata; a descriptive analysis repeated the income models among participants with prevalent recorded CKD at baseline.

SII confidence intervals were calculated using a t-distribution with three residual degrees of freedom. Aalen-Johansen estimator was used for unadjusted 5-year kidney outcome cumulative incidence, with death treated as a competing event. Associations between income and mortality within kidney function states were assessed using Cox models adjusted for the primary covariates, and an income-by-kidney-state interaction was tested using a likelihood-ratio test. Standardized 3-year mortality risks were estimated separately for each kidney state using Kaplan-Meier-based infinitesimal-jackknife pseudo-observations and identity-link regression adjusted for the primary covariates. HC1 robust standard errors were used to calculate Wald confidence intervals, without restricting their limits to the 0%-100% range. Absolute risk differences were calculated by subtracting the estimated risk in the highest-income group from that in the lowest-income group. Differences in risk differences across kidney states were assessed using eGFR ≥60 mL/min/1.73 m² as the reference, with variances summed from the separate, nonoverlapping state-specific models. These cross-state comparisons were considered descriptive. A pooled Cox model with income-by-state interaction provided standardized 2- and 3-year risks as a sensitivity analysis, with percentile intervals from 300 participant-level bootstrap replicates. Other sensitivity analyses changed mortality censoring, kidney-failure chronicity requirements, or eligibility based on numeric baseline eGFR.

## Results

### Study Population

Among 481,849 individuals who met the age and follow-up criteria, 218,098 were included in the study. Exclusions were 207,335 individuals whose baseline kidney state could not be determined, 54,482 without a household income response, and 1,934 without an education response (Supplementary Table S1; Supplementary Figure S9). Mean age was 54.3 years, and 132,327 individuals (60.7%) were female. Median follow-up was 2.76 years for kidney outcomes, and 3.62 years for mortality. Compared with individuals in the highest income category, those in the lowest category were younger, had lower educational attainment, and more often lived in deprived neighborhoods (Table 1). Baseline characteristics by educational attainment and by kidney state are shown in Supplementary Tables S2 and S3, and the availability of the neighborhood deprivation index in Supplementary Table S4. At baseline, 2,440 individuals had kidney failure, and 1,955 had received a kidney transplant.

**Table 1.**
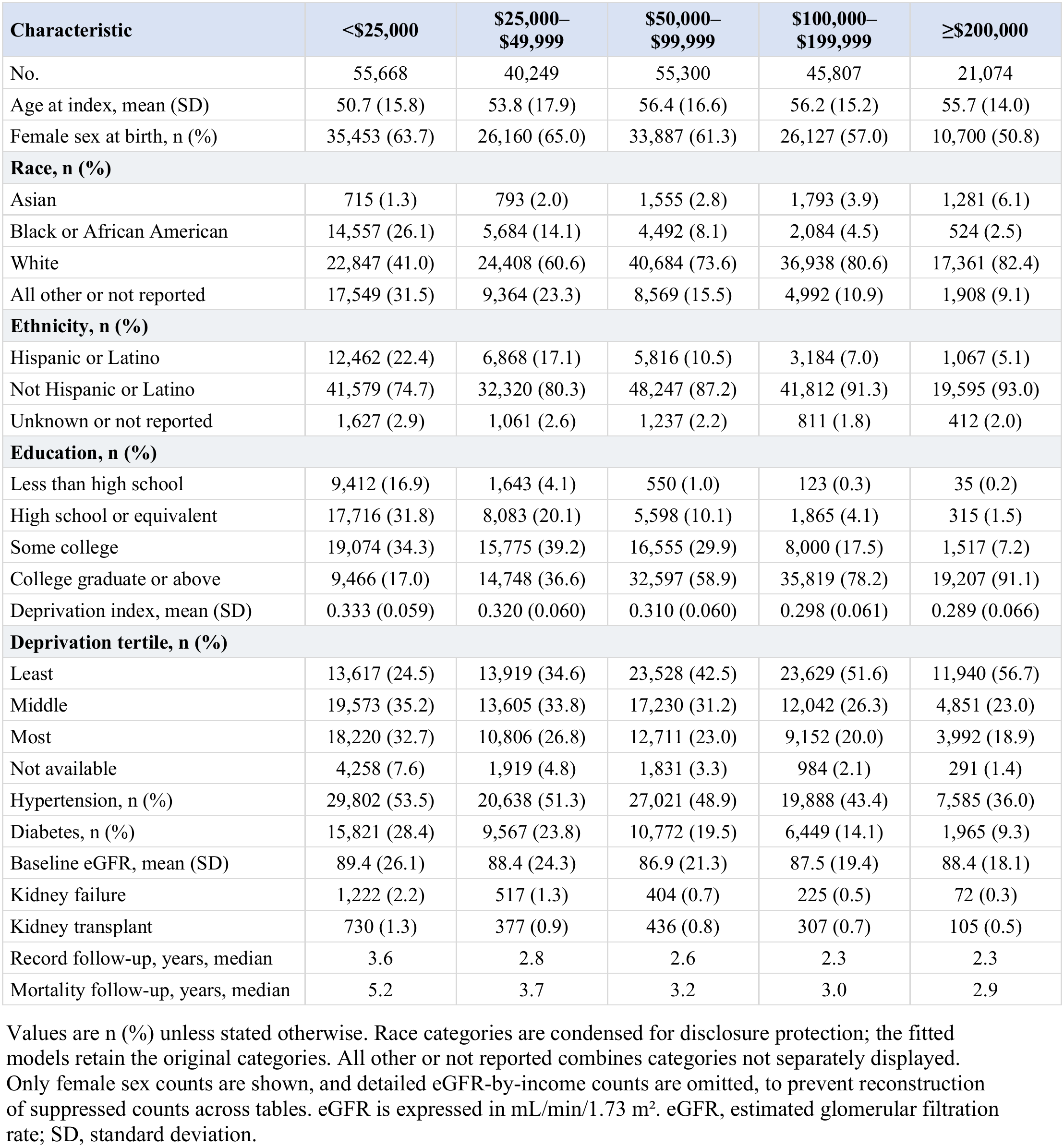
Baseline characteristics by household income.

| Characteristic | <\$25,000 | \$25,000–\$49,999 | \$50,000–\$99,999 | \$100,000–\$199,999 | ≥\$200,000 |
|---|---|---|---|---|---|
| No. | 55,668 | 40,249 | 55,300 | 45,807 | 21,074 |
| Age at index, mean (SD) | 50.7 (15.8) | 53.8 (17.9) | 56.4 (16.6) | 56.2 (15.2) | 55.7 (14.0) |
| Female sex at birth, n (%) | 35,453 (63.7) | 26,160 (65.0) | 33,887 (61.3) | 26,127 (57.0) | 10,700 (50.8) |
| **Race, n (%)** |  |  |  |  |  |
| Asian | 715 (1.3) | 793 (2.0) | 1,555 (2.8) | 1,793 (3.9) | 1,281 (6.1) |
| Black or African American | 14,557 (26.1) | 5,684 (14.1) | 4,492 (8.1) | 2,084 (4.5) | 524 (2.5) |
| White | 22,847 (41.0) | 24,408 (60.6) | 40,684 (73.6) | 36,938 (80.6) | 17,361 (82.4) |
| All other or not reported | 17,549 (31.5) | 9,364 (23.3) | 8,569 (15.5) | 4,992 (10.9) | 1,908 (9.1) |
| **Ethnicity, n (%)** |  |  |  |  |  |
| Hispanic or Latino | 12,462 (22.4) | 6,868 (17.1) | 5,816 (10.5) | 3,184 (7.0) | 1,067 (5.1) |
| Not Hispanic or Latino | 41,579 (74.7) | 32,320 (80.3) | 48,247 (87.2) | 41,812 (91.3) | 19,595 (93.0) |
| Unknown or not reported | 1,627 (2.9) | 1,061 (2.6) | 1,237 (2.2) | 811 (1.8) | 412 (2.0) |
| **Education, n (%)** |  |  |  |  |  |
| Less than high school | 9,412 (16.9) | 1,643 (4.1) | 550 (1.0) | 123 (0.3) | 35 (0.2) |
| High school or equivalent | 17,716 (31.8) | 8,083 (20.1) | 5,598 (10.1) | 1,865 (4.1) | 315 (1.5) |
| Some college | 19,074 (34.3) | 15,775 (39.2) | 16,555 (29.9) | 8,000 (17.5) | 1,517 (7.2) |
| College graduate or above | 9,466 (17.0) | 14,748 (36.6) | 32,597 (58.9) | 35,819 (78.2) | 19,207 (91.1) |
| Deprivation index, mean (SD) | 0.333 (0.059) | 0.320 (0.060) | 0.310 (0.060) | 0.298 (0.061) | 0.289 (0.066) |
| **Deprivation tertile, n (%)** |  |  |  |  |  |
| Least | 13,617 (24.5) | 13,919 (34.6) | 23,528 (42.5) | 23,629 (51.6) | 11,940 (56.7) |
| Middle | 19,573 (35.2) | 13,605 (33.8) | 17,230 (31.2) | 12,042 (26.3) | 4,851 (23.0) |
| Most | 18,220 (32.7) | 10,806 (26.8) | 12,711 (23.0) | 9,152 (20.0) | 3,992 (18.9) |
| Not available | 4,258 (7.6) | 1,919 (4.8) | 1,831 (3.3) | 984 (2.1) | 291 (1.4) |
| Hypertension, n (%) | 29,802 (53.5) | 20,638 (51.3) | 27,021 (48.9) | 19,888 (43.4) | 7,585 (36.0) |
| Diabetes, n (%) | 15,821 (28.4) | 9,567 (23.8) | 10,772 (19.5) | 6,449 (14.1) | 1,965 (9.3) |
| Baseline eGFR, mean (SD) | 89.4 (26.1) | 88.4 (24.3) | 86.9 (21.3) | 87.5 (19.4) | 88.4 (18.1) |
| Kidney failure | 1,222 (2.2) | 517 (1.3) | 404 (0.7) | 225 (0.5) | 72 (0.3) |
| Kidney transplant | 730 (1.3) | 377 (0.9) | 436 (0.8) | 307 (0.7) | 105 (0.5) |
| Record follow-up, years, median | 3.6 | 2.8 | 2.6 | 2.3 | 2.3 |
| Mortality follow-up, years, median | 5.2 | 3.7 | 3.2 | 3.0 | 2.9 |

Values are n (%) unless stated otherwise. Race categories are condensed for disclosure protection; the fitted models retain the original categories. All other or not reported combines categories not separately displayed. Only female sex counts are shown, and detailed eGFR-by-income counts are omitted, to prevent reconstruction of suppressed counts across tables. eGFR is expressed in mL/min/1.73 m<sup>2</sup>. eGFR, estimated glomerular filtration rate; SD, standard deviation.

### Socioeconomic position, mortality, and kidney outcomes

During follow-up, 4,777 individuals died. Among the 209,916 individuals included in the kidney-function decline analysis, 6,423 experienced a sustained 40% decline in eGFR. Compared with participants with household income of $200,000 or more, those with income below $25,000 had higher hazards of mortality (adjusted HR, 2.68; 95% CI, 2.31-3.12) and sustained kidney function decline (adjusted HR, 3.57; 95% CI, 3.08-4.13).

Among 194,814 individuals included in the recorded CKD analyses, 7,870 had a first recorded CKD diagnosis and 4,716 had a first recorded diagnosis of CKD category G3a or worse. Kidney failure occurred in 1,799 of 209,916 eligible individuals. Comparing the lowest with the highest income category, adjusted HRs were 2.13 (95% CI, 1.90–2.38) for recorded CKD diagnosis, 2.15 (95% CI, 1.86–2.49) for recorded CKD category G3a or worse, and 3.08 (95% CI, 2.30–4.11) for kidney failure (Table 2; Figure 1).

**Figure 1.**
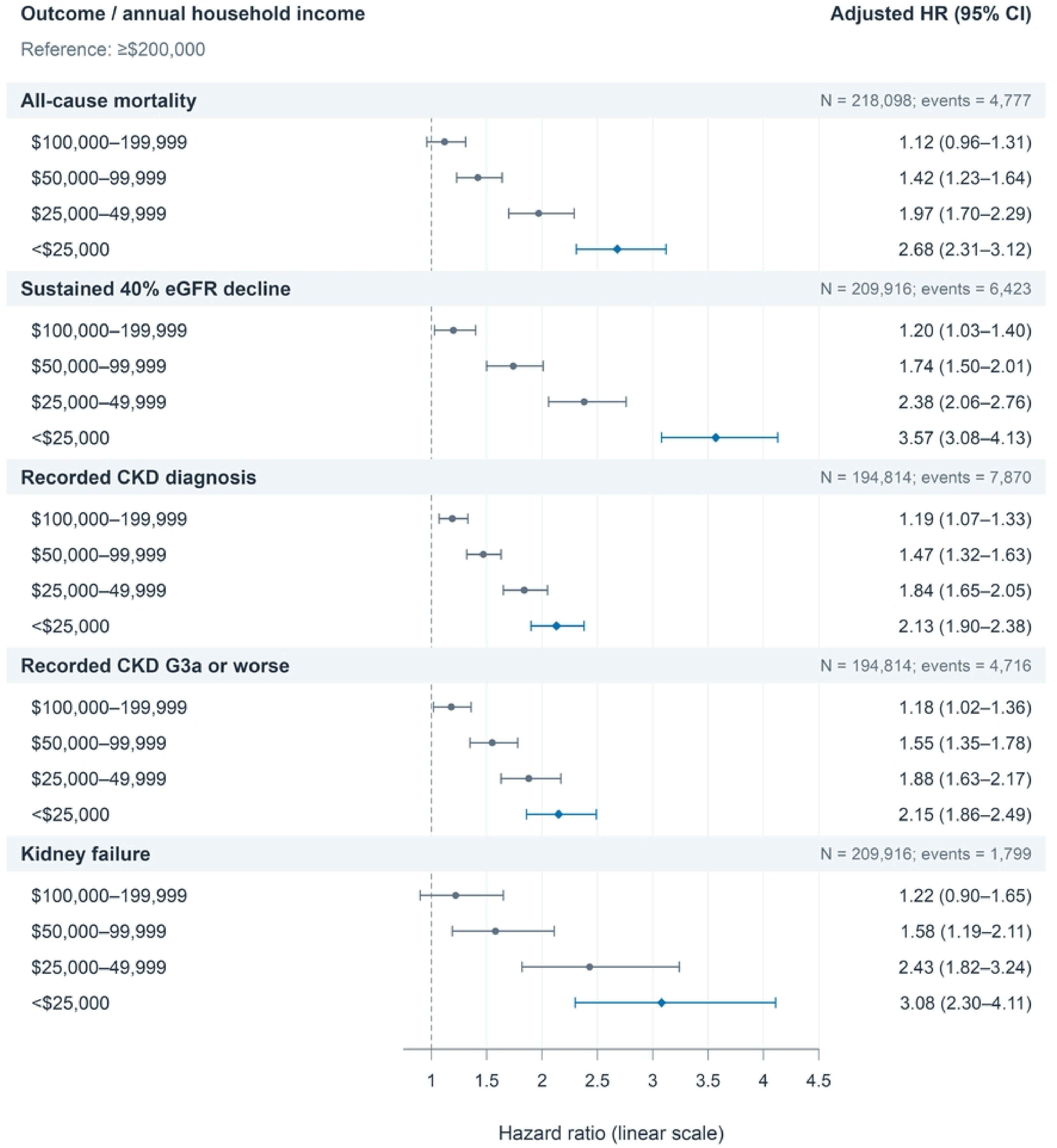
Household income and mortality and kidney outcomes. Adjusted hazard ratios and 95% confidence intervals compare each household income category with income of $200,000 or more for all-cause mortality, sustained 40% eGFR decline, newly recorded CKD diagnosis, newly recorded CKD category G3a or worse, and kidney failure. Models include income, education, and neighborhood deprivation together and are adjusted for age, sex, race, and ethnicity; each outcome has its own risk set, and the number of participants and events is shown for each. The hazard-ratio axis is linear; blue diamonds mark income below $25,000. CI indicates confidence interval; CKD, chronic kidney disease; eGFR, estimated glomerular filtration rate; HR, hazard ratio.

**Table 2.** Incidence rates and adjusted hazard ratios by household income.

| **Household income** | **No.** | **Events** | **Person-years** | **Rate\*** | **Adjusted HR<br>(95% CI)** | **P** |
|---|---|---|---|---|---|---|
| **All-cause mortality** |  |  |  |  |  |  |
| ≥\$200,000 | 21,074 | 229 | 71,759 | 3.19 | 1 (reference) | — |
| \$100,000–\$199,999 | 45,807 | 585 | 156,009 | 3.75 | 1.12 (0.96-1.31) | 0.13 |
| \$50,000–\$99,999 | 55,300 | 1,004 | 196,621 | 5.11 | 1.42 (1.23-1.64) | <0.001 |
| \$25,000–\$49,999 | 40,249 | 1,042 | 153,159 | 6.80 | 1.97 (1.70-2.29) | <0.001 |
| <\$25,000 | 55,668 | 1,917 | 243,753 | 7.86 | 2.68 (2.31-3.12) | <0.001 |
| **Sustained 40% eGFR decline** |  |  |  |  |  |  |
| ≥\$200,000 | 20,508 | 222 | 59,547 | 3.73 | 1 (reference) | — |
| \$100,000–\$199,999 | 44,493 | 613 | 127,698 | 4.80 | 1.20 (1.03-1.40) | 0.02 |
| \$50,000–\$99,999 | 53,528 | 1,227 | 158,223 | 7.75 | 1.74 (1.50-2.01) | <0.001 |
| \$25,000–\$49,999 | 38,670 | 1,321 | 118,536 | 11.14 | 2.38 (2.06-2.76) | <0.001 |
| <\$25,000 | 52,717 | 3,040 | 177,119 | 17.16 | 3.57 (3.08-4.13) | <0.001 |
| **Newly recorded CKD diagnosis** |  |  |  |  |  |  |
| ≥\$200,000 | 19,704 | 437 | 56,570 | 7.72 | 1 (reference) | — |
| \$100,000–\$199,999 | 42,017 | 1,158 | 119,141 | 9.72 | 1.19 (1.07-1.33) | 0.002 |
| \$50,000–\$99,999 | 49,656 | 1,902 | 145,005 | 13.12 | 1.47 (1.32-1.63) | <0.001 |
| \$25,000–\$49,999 | 35,385 | 1,711 | 107,330 | 15.94 | 1.84 (1.65-2.05) | <0.001 |
| <\$25,000 | 48,052 | 2,662 | 162,257 | 16.41 | 2.13 (1.90-2.38) | <0.001 |
| **Newly recorded CKD category G3a or worse** |  |  |  |  |  |  |
| ≥\$200,000 | 19,704 | 258 | 56,966 | 4.53 | 1 (reference) | — |
| \$100,000–\$199,999 | 42,017 | 681 | 120,223 | 5.66 | 1.18 (1.02-1.36) | 0.03 |
| \$50,000–\$99,999 | 49,656 | 1,215 | 146,683 | 8.28 | 1.55 (1.35-1.78) | <0.001 |
| \$25,000–\$49,999 | 35,385 | 1,049 | 109,009 | 9.62 | 1.88 (1.63-2.17) | <0.001 |
| <\$25,000 | 48,052 | 1,513 | 165,445 | 9.15 | 2.15 (1.86-2.49) | <0.001 |
| **Kidney failure** |  |  |  |  |  |  |
| ≥\$200,000 | 20,508 | 57 | 59,931 | 0.95 | 1 (reference) | — |
| \$100,000–\$199,999 | 44,493 | 161 | 128,848 | 1.25 | 1.22 (0.90-1.65) | 0.20 |
| \$50,000–\$99,999 | 53,528 | 301 | 160,686 | 1.87 | 1.58 (1.19-2.11) | 0.002 |
| \$25,000–\$49,999 | 38,670 | 395 | 121,232 | 3.26 | 2.43 (1.82-3.24) | <0.001 |
| <\$25,000 | 52,717 | 885 | 183,905 | 4.81 | 3.08 (2.30-4.11) | <0.001 |

Rates are crude events per 1,000 person-years. Models include household income, education, neighborhood deprivation, linear age, sex at birth, race, and ethnicity. Household income ≥\$200,000 is the reference category. Each outcome has its own risk set. CI, confidence interval; CKD, chronic kidney disease; eGFR, estimated glomerular filtration rate; HR, hazard ratio.

Among individuals who had not completed high school, the adjusted HR for mortality was 1.41 (95% CI, 1.25-1.60) compared with college graduates. The corresponding adjusted hazard ratio for sustained eGFR decline was 1.42 (95% CI, 1.29-1.58). Both models were adjusted for household income, neighborhood deprivation, and demographic covariates. Individuals living in the most deprived neighborhood tertile had higher hazards of all four kidney outcomes than those in the least deprived tertile. Among the four kidney outcomes, the HR was lowest for sustained eGFR decline at 1.09 (95% CI, 1.02-1.17) and highest for kidney failure at 1.42 (95% CI, 1.25-1.61). No association with mortality was observed for this comparison (HR, 1.00; 95% CI, 0.93-1.08) (Supplementary Table S5; Supplementary Figure S1).

### Income inequalities across outcomes

Inequality was greater for sustained eGFR decline and kidney failure than for outcomes based on recorded CKD diagnoses. The relative index of inequality was 6.84 (95% CI, 6.15–7.61) for sustained eGFR decline and 6.06 (95% CI, 4.94–7.44) for kidney failure. Lower estimates were observed for recorded CKD diagnosis (3.02; 95% CI, 2.76–3.30) and CKD category G3a or worse (2.94; 95% CI, 2.62–3.31). For mortality, the relative index of inequality was 4.70 (95% CI, 4.17–5.29). Sustained eGFR decline also showed the largest crude absolute inequality, with a slope index of 16.98 additional events per 1,000 person-years (95% CI, 10.27–23.70). Differences in inequality estimates across outcomes were described but not formally tested (Supplementary Table S5; Supplementary Figure S10).

### Mortality by baseline kidney state

Among individuals with baseline eGFR ≥60 mL/min/1.73 m², those in the lowest income category had a higher risk of mortality than those in the highest category (adjusted HR, 2.73; 95% CI, 2.28–3.26). The corresponding HR was 1.86 (95% CI, 1.32–2.61) for eGFR 30–59 mL/min/1.73 m², 1.24 (95% CI, 0.50–3.05) for eGFR <30 mL/min/1.73 m², 1.31 (95% CI, 0.58–2.94) for kidney failure, and 1.71 (95% CI, 0.70–4.16) for kidney transplantation. Estimates were less precise in the latter three groups, with wide confidence intervals. The overall interaction between income and baseline kidney state was nominally significant (likelihood ratio χ² = 26.5; 16 degrees of freedom; P = 0.047), although the state-specific estimates were imprecise (Table 3).

**Table 3.** Mortality by baseline kidney state: relative and standardized 3-year absolute associations.

| A. Adjusted mortality hazard ratios |  |  |  |  |  |
| Baseline kidney state | No. | Deaths | Adjusted HR (95% CI) |  |  |
|---|---|---|---|---|---|
| eGFR ≥60 | 193,804 | 3,099 | 2.73 (2.28-3.26) |  |  |
| eGFR 30-59 | 18,319 | 1,007 | 1.86 (1.32-2.61) |  |  |
| eGFR <30 | 1,580 | 184 | 1.24 (0.50-3.05) |  |  |
| Kidney failure | 2,440 | 294 | 1.31 (0.58-2.94) |  |  |
| Kidney transplant | 1,955 | 193 | 1.71 (0.70-4.16) |  |  |
| **B. Standardized 3-year mortality risks and absolute differences** |  |  |  |  |  |
| **Baseline kidney state** | **Low-income risk, % (95% CI)** | **High-income risk, % (95% CI)** | **Risk difference, pp (95% CI)** | **Difference in risk differences, pp (95% CI)** | **P** |
| eGFR ≥60 | 1.77 (1.61 to 1.92) | 0.73 (0.58 to 0.87) | 1.04 (0.81 to 1.27) | Reference | — |
| eGFR 30-59 | 5.09 (4.30 to 5.87) | 3.21 (2.08 to 4.34) | 1.88 (0.44 to 3.33) | 0.84 (-0.62 to 2.30) | 0.261 |
| eGFR <30 | 10.93 (8.09 to 13.78) | 3.43 (-2.89 to 9.75) | 7.51 (0.16 to 14.85) | 6.47 (-0.88 to 13.81) | 0.085 |
| Kidney failure | 11.09 (8.98 to 13.19) | 8.99 (1.87 to 16.10) | 2.10 (-5.50 to 9.70) | 1.06 (-6.55 to 8.66) | 0.786 |
| Kidney transplant | 7.14 (5.01 to 9.27) | 4.94 (1.12 to 8.76) | 2.20 (-2.33 to 6.73) | 1.15 (-3.38 to 5.69) | 0.618 |

Low income denotes household income <\$25,000; high income denotes ≥\$200,000. Models are adjusted for age, sex, race, ethnicity, education, and neighborhood deprivation. Panel A participant and death counts describe the full state-specific mortality risk sets and available follow-up. Panel B uses separate kidney-state pseudo-observation models, each standardized over that state's participants, with HC1 standard errors for 95% confidence intervals. Cross-state comparisons are descriptive. Risk differences are low minus high income. Differences in risk differences use eGFR ≥60 as the reference; P values in panel B refer to these comparisons. Mortality-risk confidence intervals are not truncated at 0% or 100%. eGFR is expressed in mL/min/1.73 m<sup>2</sup>. CI, confidence interval; eGFR, estimated glomerular filtration rate; HR, hazard ratio; pp, percentage points.

Among individuals with eGFR ≥60 mL/min/1.73 m², standardized 3-year mortality risks were 1.77% in the lowest income category and 0.73% in the highest, a difference of 1.04 percentage points (95% CI, 0.81–1.27). Among those with eGFR 30–59, risks were 5.09% and 3.21%, respectively, a difference of 1.88 percentage points (95% CI, 0.44–3.33). Risk differences were 7.51 percentage points (95% CI, 0.16–14.85) for eGFR <30, 2.10 percentage points (95% CI, −5.50 to 9.70) for kidney failure, and 2.20 percentage points (95% CI, −2.33 to 6.73) for kidney transplantation. None of these risk differences differed significantly from that in the eGFR ≥60 group (all P ≥0.05) (Table 3; Figure 2).

**Figure 2.**
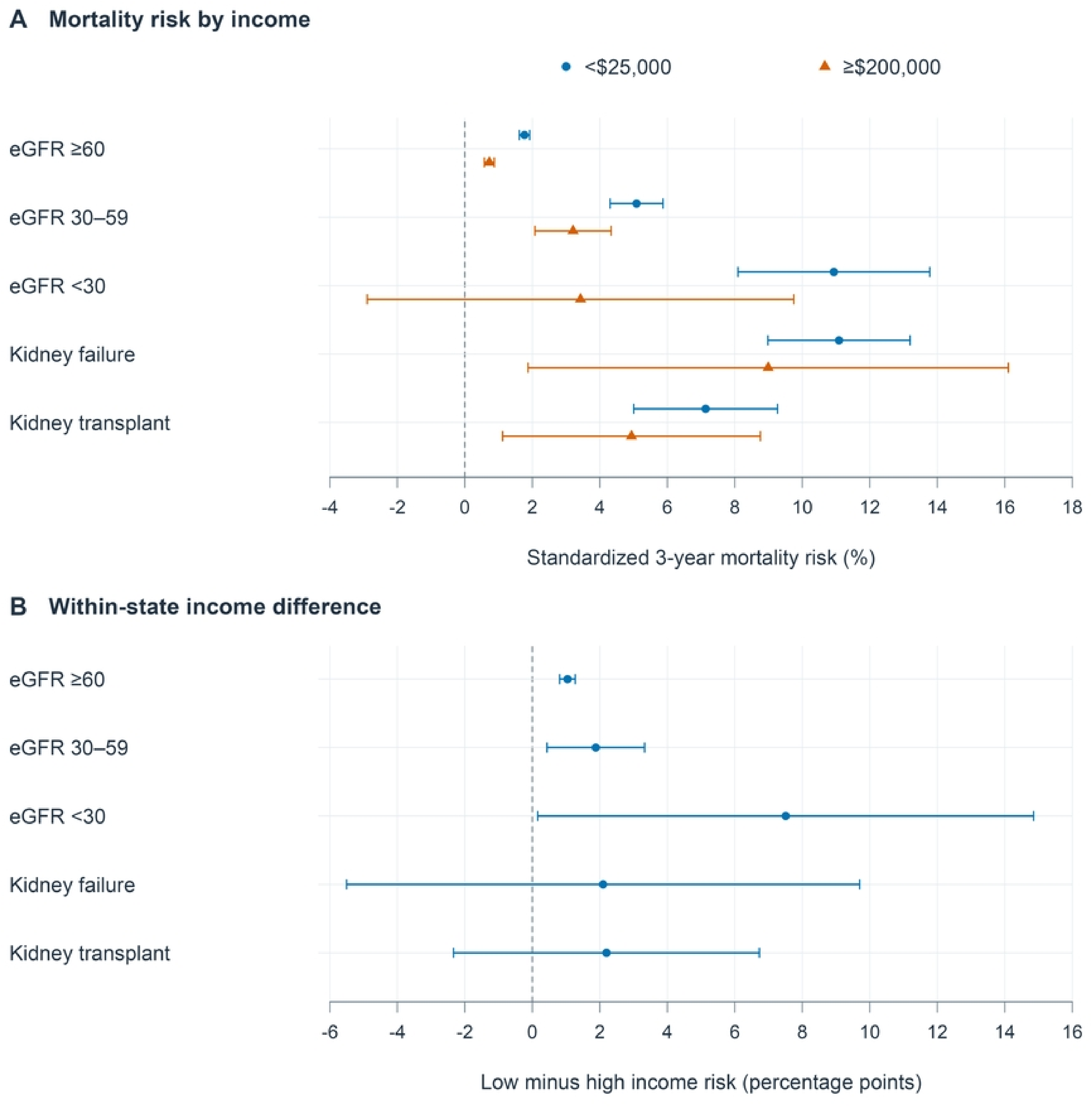
Standardized 3-year mortality risks by household income within baseline kidney states. (A) Standardized 3-year mortality risks for household income below $25,000 (blue circles) and $200,000 or more (orange triangles). (B) Low-minus-high standardized risk differences in percentage points. Estimates are from separate kidney-state pseudo-observation regressions with HC1 heteroskedasticity-consistent standard errors, adjusted for education, neighborhood deprivation, age, sex, race, and ethnicity, each standardized over that state’s participants; cross-state comparisons are descriptive. Confidence intervals are not truncated at 0% or 100%; bounds outside this range reflect sparse support and model uncertainty. Points are estimates and bars are 95% confidence intervals. CI indicates confidence interval; eGFR, estimated glomerular filtration rate.

### Sensitivity and exploratory analyses

The proportional hazards assumption was not met for mortality, sustained eGFR decline, or either recorded CKD outcome. However, the association between lower income and higher hazards remained during both the first 2 years and subsequent follow-up. For mortality, the HR comparing the lowest with the highest income category was 2.18 (95% CI, 1.74-2.73) during the first two years and 3.12 (95% CI, 2.55-3.81) thereafter. The corresponding HRs for sustained eGFR decline were 4.21 (95% CI, 3.42-5.18) and 2.95 (95% CI, 2.40-3.64), respectively (Supplementary Table S8).

With death treated as a competing event, the unadjusted 5-year cumulative incidence of all kidney outcomes was higher in the lowest income category than in the highest. The 5-year cumulative incidence in the lowest and highest income categories was 8.00% and 1.95%, respectively, for sustained eGFR decline; 7.49% and 3.84% for recorded CKD diagnosis; 4.33% versus 2.23% for recorded CKD category G3a or worse; and 2.32% versus 0.46% for kidney failure (Supplementary Table S7; Supplementary Figure S2).

In sensitivity analyses, censoring mortality follow-up at the last recorded EHR contact yielded an HR of 2.95 (95% CI, 2.38–3.65), whereas the primary analysis included 2,256 deaths occurring after the last EHR contact. Restricting the analysis to individuals with a numeric baseline eGFR yielded a mortality HR of 2.70 (95% CI, 2.32–3.14). When the chronicity requirements for kidney failure were removed, the number of events increased to 2,564, with an HR of 2.78 (95% CI, 2.21–3.51) (Supplementary Table S10; Supplementary Figure S4). In the pooled Cox sensitivity analysis, which assumes proportional hazards that the diagnostics rejected, differences in standardized risk differences versus eGFR ≥60 mL/min/1.73 m² were larger and their percentile intervals excluded zero for eGFR 30–59 (1.43 percentage points; 95% CI, 0.17–2.46) and kidney transplantation (3.95 percentage points; 95% CI, 0.52–6.93) at 3 years (Supplementary Table S12; Supplementary Figure S8). Exploratory subgroup analyses are shown in Supplementary Table S9 and Supplementary Figure S3, categorical associations within sex, race, and deprivation strata in Supplementary Table S11 and Supplementary Figures S5 to S7, and income associations among participants with prevalent recorded CKD in Supplementary Table S6.

## Discussion

Our analysis of this large-scale observational All of Us cohort revealed that lower income was associated with higher mortality and all four kidney outcomes. In particular, income-gradient estimates were numerically larger for sustained kidney function decline and kidney failure than for a first recorded CKD diagnosis, with substantial differences also evident in the 5-year cumulative incidence of kidney outcomes. Education and neighborhood deprivation demonstrated different patterns of association, highlighting the importance of considering individual and neighborhood socioeconomic circumstances together. Interestingly, income-related mortality hazard ratios were numerically lower in more advanced baseline kidney states. Income associations also varied over follow-up, with mortality hazard ratios numerically larger later and hazard ratios for sustained kidney function decline larger earlier.

Previous large-scale cohort studies have already linked socioeconomic disadvantage to kidney function decline, mortality, and kidney failure. In a nationwide Korean cohort of 7,405,715 adults with a median follow-up duration of 4.8 years, individuals in the lowest income decile had a higher risk of incident CKD than those in the sixth decile (adjusted HR, 1.30; 95% CI, 1.26– 1.33) (12). The association was similar when incident CKD required both a ≥25% decline in eGFR and an eGFR below 60 mL/min/1.73 m² (adjusted HR, 1.32; 95% CI, 1.27–1.37).

Similarly, among 1,405,016 English primary-care patients followed for a median of 7.5 years, those living in the most deprived 20% of areas had higher hazards of CKD stages 4–5 and ESRD than those in the least deprived 20% (age- and sex-adjusted HRs, 1.76; 95% CI, 1.68–1.84 and 1.82; 95% CI, 1.56–2.12, respectively) (13). Furthermore, among 2,761 individuals with prevalent CKD stage 3 or 4, annual household income below $20,000 was associated with higher all-cause mortality than income of $20,000 or more (adjusted HR, 1.58; 95% CI, 1.24-2.00) (14). Our study further extends prior findings in a large, ethnically diverse U.S. cohort with linked longitudinal electronic health records by jointly examining income, education, and neighborhood deprivation in relation to sustained kidney function decline, recorded CKD, kidney failure, and mortality, while quantifying 5-year kidney-event incidence across income groups and evaluating mortality inequalities across baseline kidney states.

Importantly, the income gradients were accompanied by substantial differences in 5-year kidney outcomes. At 5 years, estimated cumulative incidence of sustained kidney function decline was 8.00% in the lowest-income group, compared with 1.95% in the highest-income group. The corresponding estimates for kidney failure were 2.32% and 0.46%. These findings are consistent with previous U.S. studies showing greater burden of kidney failure among individuals with lower incomes and in counties with greater poverty. Among 79,943 individuals in the Southern Community Cohort Study, annual household income below $15,000 was associated with a higher risk of kidney failure than income of $15,000 or more (adjusted HR, 1.5; 95% CI, 1.2-1.8) (15). Similarly, a national U.S. analysis also found that the excess incidence of kidney failure in high-poverty compared with low-poverty counties increased from 42.8 cases per million in 2000-2005 to 100.1 cases per million in 2012-2017, despite an overall national decline in kidney failure incidence (16). Our study extends these reports on kidney failure by also quantifying income inequalities in sustained kidney function decline confirmed by repeated measurements. Incorporating all income categories demonstrated that sustained kidney function decline had numerically the largest relative and absolute income inequalities (RII, 6.84; SII, 16.98 events per 1,000 person-years).

Furthermore, education and neighborhood deprivation demonstrated different patterns of associations across outcomes. Lower education was associated with higher risks of mortality and sustained kidney function decline. On the other hand, greater neighborhood deprivation was associated with adverse kidney outcomes despite demonstrating no clear associations with mortality. These findings highlight the importance of considering both individual and neighborhood socioeconomic circumstances when examining inequalities across kidney outcomes. Among 5,095 individuals in the German CKD cohort with a follow-up duration of 6.5 years, lower education was associated with higher mortality (age- and sex-adjusted HR, 1.48; 95% CI, 1.16–1.90) and kidney failure requiring dialysis (HR, 1.54; 95% CI, 1.15–2.05) compared with higher education (17). Nevertheless, these associations were no longer statistically significant after further adjustment for kidney disease severity and potential mediators, including income. In the Cardiovascular Health Study of 4,735 individuals aged 65 years or older, those living in neighborhoods with the lowest socioeconomic status had a 50% higher risk of progressive CKD compared to those in the highest socioeconomic quartile, even after adjustment for demographic characteristics, baseline creatinine, and individual-level socioeconomic factors (18). This association persisted after further adjustment for lifestyle factors, diabetes, and hypertension, whereas individual income and education were not independently associated with progressive CKD after accounting for the other SES measures. In our broader large-scale U.S. adult cohort, lower education remained associated with mortality and sustained kidney function decline after accounting for income and neighborhood deprivation. These findings suggest that income alone may not fully capture socioeconomic inequalities across kidney outcomes.

Moreover, relative income associations with mortality were numerically smaller among individuals with more advanced kidney disease. Specifically, the adjusted mortality hazard ratio comparing the lowest- with the highest-income group was 2.73 (95% CI, 2.28-3.26) among participants with baseline eGFR ≥60 mL/min/1.73 m², compared with 1.24 (95% CI, 0.50–3.05) among those with eGFR <30 mL/min/1.73 m². Nevertheless, evidence of heterogeneity across kidney-function strata was weak, and estimates among participants with advanced kidney dysfunction were imprecise. Furthermore, the standardized 3-year analysis did not establish differences in absolute mortality gaps across states, although the intervals were wide and the pooled Cox sensitivity analysis, which depends on a rejected proportionality assumption, suggested larger gaps in two states. A similar numerical pattern was reported in a Grampian population study. The adjusted mortality hazard ratio comparing residents of the most deprived areas with those in all other areas was 1.21 (95% CI, 1.14–1.28) among people presenting with eGFR <60 mL/min/1.73 m² and 1.11 (95% CI, 1.04–1.19) among those presenting with eGFR <30 mL/min/1.73 m² (3). Our study complements this evidence by estimating standardized absolute differences in three-year mortality between income groups across baseline kidney states.

Income associations also varied over follow-up. Comparing the lowest-with the highest-income group, the adjusted hazard ratio for mortality was 2.18 during the first two years of follow-up and 3.12 thereafter. The corresponding hazard ratios for sustained kidney function decline were 4.21 and 2.95. Although the strengths of these associations varied over time, income-related inequalities remained evident in both early and later follow-up. These findings suggest that a single overall hazard ratio may obscure changes in the magnitude of income associations during follow-up. Differences in participants’ health at baseline or their use of healthcare potentially could explain these patterns; nevertheless, we did not test those explanations. Future studies could potentially clarify why the associations varied over time.

Although our large-scale observational cohort study provides valuable insights into socioeconomic inequalities across kidney outcomes and mortality, several limitations should be acknowledged. First, given the observational nature of the study, the observed associations should not be interpreted as causal relationships. Second, socioeconomic circumstances were assessed only at baseline, so subsequent income changes were not captured, while neighborhood deprivation measured using three-digit ZIP codes may obscure local differences. Third, incomplete EHR coverage and differences in testing and coding may have affected outcome detection. Incident CKD was identified from diagnosis records and a laboratory definition based on repeated low eGFR values was not applied; recorded creatinine units were not verified; and deaths near the end of the data period may be incompletely captured because of reporting lag.

Some creatinine measurements used to assess baseline kidney function were obtained after the socioeconomic survey, creating uncertainty about kidney status at the time of income assessment. Fourth, only 218,098 of 481,849 otherwise eligible participants had a determined kidney state and complete socioeconomic responses, so selection related to income, illness, and healthcare use is possible. Finally, as the All of Us Research Program is a volunteer-based cohort, participants may differ from the general population, potentially limiting the generalizability of our findings. Therefore, further studies with repeated socioeconomic assessments, more detailed neighborhood information, and standardized kidney function measurements are needed to clarify these associations and identify potential ways to reduce socioeconomic inequalities in kidney health.

## Acknowledgements

We gratefully acknowledge All of Us participants for their contributions, without whom this research would not have been possible. We also thank the National Institutes of Health’s All of Us Research Program for making available the participant data examined in this study.

## Conflict of Interest Statement

The authors declare no competing interests.

## Authors’ Contributions

A.D.: Conceptualization, methodology, formal analysis, validation, and writing-review and editing. I.G.: Conceptualization, methodology, formal analysis, validation, writing original draft, and writing-review and editing. M.A.E.: Writing-original draft. M.B.: Conceptualization and writing-review and editing. S.C.: Conceptualization, supervision, visualization, and writing-review and editing. F.M., C.Z., and M.K.: Conceptualization, methodology, validation, supervision, and writing-review and editing.

## Data Availability Statement

This study used the All of Us Research Program Controlled Tier Dataset C2025Q4R6, available to authorized users through the Researcher Workbench under the program Data Use and Registration Agreement. Participant-level data cannot be distributed by the authors. Disclosure-safe aggregate results and ordered analysis code are intended for public release in a versioned repository. The analysis code is maintained at https://github.com/atalaydemiray/aou-ses-kidney-analysis.

## Supporting information

S1 Appendix. Supplementary Methods, Supplementary Tables S1 to S12, and Supplementary Figures S1 to S10.

S1 Checklist. STROBE checklist for cohort studies.

## Notes

### Competing Interest Statement

The authors have declared no competing interest.

### Author Declarations

All of Us Institutional Review Board, protocol 11-01139 (program protocol and participant consent) Yale University Institutional Review Board, protocol 2000043320 (determination of not human subjects research, 14 July 2026).

## References

1. Braveman P, Gruskin S. Defining equity in health. J Epidemiol Community Health. 2003;57(4):254–8.

2. World Health Organization. Universal health coverage (UHC) 2025 [Available from: https://www.who.int/news-room/fact-sheets/detail/universal-health-coverage-%28uhc%29.

3. Sawhney S, Blakeman T, Blana D, Boyers D, Fluck N, Nath M, et al. Care processes and outcomes of deprivation across the clinical course of kidney disease: findings from a high-income country with universal healthcare. Nephrol Dial Transplant. 2023;38(5):1170–82.

4. Zeng X, Liu J, Tao S, Hong HG, Li Y, Fu P. Associations between socioeconomic status and chronic kidney disease: a meta-analysis. J Epidemiol Community Health. 2018;72(4):270–9.

5. Vart P, Grams ME, Ballew SH, Woodward M, Coresh J, Matsushita K. Socioeconomic status and risk of kidney dysfunction: the Atherosclerosis Risk in Communities study. Nephrol Dial Transplant. 2019;34(8):1361–8.

6. Ishimura N, Inoue K, Maruyama S, Nakamura S, Kondo N. Income Level and Impaired Kidney Function Among Working Adults in Japan. JAMA Health Forum. 2024;5(3):e235445.

7. Sawhney S, Atherton I, Blakeman T, Black C, Cowan E, Croucher C, et al. Individual and neighborhood-level social and deprivation factors impact kidney health in the GLOMMS-CORE study. Kidney Int. 2024;106(5):928–42.

8. Denny JC, Rutter JL, Goldstein DB, Philippakis A, Smoller JW, Jenkins G, et al. The “All of Us” Research Program. N Engl J Med. 2019;381(7):668–76.

9. von Elm E, Altman DG, Egger M, Pocock SJ, Gøtzsche PC, Vandenbroucke JP. The Strengthening the Reporting of Observational Studies in Epidemiology (STROBE) statement: guidelines for reporting observational studies. PLoS Med. 2007;4(10):e296.

10. Inker LA, Eneanya ND, Coresh J, Tighiouart H, Wang D, Sang Y, et al. New Creatinine- and Cystatin C-Based Equations to Estimate GFR without Race. N Engl J Med. 2021;385(19):1737–49.

11. Moreno-Betancur M, Latouche A, Menvielle G, Kunst AE, Rey G. Relative index of inequality and slope index of inequality: a structured regression framework for estimation. Epidemiology. 2015;26(4):518–27.

12. Chang TI, Lim H, Park CH, Rhee CM, Kalantar-Zadeh K, Kang EW, et al. Association Between Income Disparities and Risk of Chronic Kidney Disease: A Nationwide Cohort Study of Seven Million Adults in Korea. Mayo Clin Proc. 2020;95(2):231–42.

13. Weldegiorgis M, Smith M, Herrington WG, Bankhead C, Woodward M. Socioeconomic disadvantage and the risk of advanced chronic kidney disease: results from a cohort study with 1.4 million participants. Nephrol Dial Transplant. 2020;35(9):1562–70.

14. Fedewa SA, McClellan WM, Judd S, Gutiérrez OM, Crews DC. The association between race and income on risk of mortality in patients with moderate chronic kidney disease. BMC Nephrol. 2014;15:136.

15. Lipworth L, Mumma MT, Cavanaugh KL, Edwards TL, Ikizler TA, Tarone RE, et al. Incidence and predictors of end stage renal disease among low-income blacks and whites. PLoS One. 2012;7(10):e48407.

16. Nguyen KH, Thorsness R, Swaminathan S, Mehrotra R, Patzer RE, Lee Y, et al. Despite National Declines In Kidney Failure Incidence, Disparities Widened Between Low- And High-Poverty US Counties. Health Aff (Millwood). 2021;40(12):1900–8.

17. Winitzki D, Zacharias HU, Nadal J, Baid-Agrawal S, Schaeffner E, Schmid M, et al. Educational Attainment Is Associated With Kidney and Cardiovascular Outcomes in the German CKD (GCKD) Cohort. Kidney Int Rep. 2022;7(5):1004–15.

18. Merkin SS, Diez Roux AV, Coresh J, Fried LF, Jackson SA, Powe NR. Individual and neighborhood socioeconomic status and progressive chronic kidney disease in an elderly population: The Cardiovascular Health Study. Soc Sci Med. 2007;65(4):809–21.

